# Engineering luminescent biosensors for point-of-care SARS-CoV-2 antibody detection

**DOI:** 10.1101/2020.08.17.20176925

**Authors:** Susanna K. Elledge, Xin X. Zhou, James R. Byrnes, Alexander J. Martinko, Irene Lui, Katarina Pance, Shion A. Lim, Jeff E. Glasgow, Anum A. Glasgow, Keirstinne Turcios, Nikita Iyer, Leonel Torres, Michael J. Peluso, Timothy J. Henrich, Taia T. Wang, Cristina M. Tato, Kevin K. Leung, Bryan Greenhouse, James A. Wells

## Abstract

Current serology tests for SARS-CoV-2 antibodies mainly take the form of enzyme-linked immunosorbent assays or lateral flow assays, with the former being laborious and the latter being expensive and often lacking sufficient sensitivity and scalability. Here we present the development and validation of a rapid, low-cost solution-based assay to detect antibodies in serum, plasma, whole blood, and saliva, using rationally designed split luciferase antibody biosensors (spLUC). This new assay, which generates quantitative results in as short as 5 minutes, substantially reduces the complexity and improves the scalability of COVID-19 antibody tests for point-of-care and broad population testing.

## INTRODUCTION

As the COVD-19 pandemic continues worldwide, broad testing for SARS-CoV-2 infection still faces severe limitations. While nucleic acid testing is critical to detecting the virus, serological antibody tests are vital tools for monitoring the dynamic human humoral response to SARS-CoV-2 viral infection and vaccines (Krammer and Simon, 2020). Antibody tests serve as a complement or an alternative to nucleic acid diagnostics for patients with a low viral load or for low-resource areas where expensive RT-PCR testing is difficult to access (Long et al., 2020; To et al., 2020; Zhao et al., 2020). Serological tests also support therapeutic development either through identification of individuals who could serve as donors for convalescent serum therapeutics (Casadevall and Pirofski, 2020), or patients with potentially strong neutralizing antibodies that can be produced *in vitro* as new antivirals and prophylactics (Robbiani et al., 2020; Rogers et al., 2020). Importantly, as a vaccine is developed, population-scale, longitudinal evaluation of antibody responses is needed to determine the response to vaccination and the strength and duration of immunity. This would be greatly accelerated with an assay that is simple, rapid, and high-throughput without sacrificing accuracy and sensitivity (Lynch et al., 2020; Okba et al., 2020; Seow et al., 2020; Smith et al., 2020; Yu et al., 2020; van Doremalen et al., 2020).

Traditional serological assays are not optimal in the face of this broad pandemic. The most widely used laboratory serological tests take the form of enzyme-linked immunosorbent assays (ELISA) (Amanat et al., 2020; Okba et al., 2020; Tan et al., 2020b; Xiang et al., 2020), which usually entail a >2-hour protocol involving several steps of protein incubation and washes, and is not readily amenable to deployment outside of a laboratory. A faster but significantly more expensive approach is a lateral flow assay (Li et al., 2020; Lassaunière et al., 2020). However, lateral flow assays can produce less reliable results depending on the quality of the lateral flow device and different evaluation criteria (Whitman et al., 2020; Lassaunière et al., 2020). In addition, lateral flow tests poorly capture the magnitude of a patient’s antibody response as the test is qualitative and not quantitative. Here we provide a next-generation, simple, and low-cost assay to meet the mounting needs for broad antibody testing in the face of the ongoing pandemic and eventual vaccine deployment. The assay, which is compatible with serum, plasma, whole blood, and saliva samples, utilizes a simple split luciferase (spLUC) antibody sensor to generate quantitative serological data in as short as 5 minutes. Testing of over 150 patient serum/plasma samples across three validation cohorts demonstrates that the spLUC assay has both sensitivity and specificity of>98%.

## RESULTS

### Engineering split luminescent biosensors (spLUC) for SARS-CoV-2 antibody detection

When envisioning a next-generation serological assay, we hypothesized that sensitive biosensors for anti-SARS-CoV-2 antibodies could be utilized to greatly enhance the speed and simplicity of serological testing (Dixon et al., 2016). We constructed anti-SARS-CoV-2 antibody biosensors by fusing split Nanoluciferase (NanoLuc) fragments SmBiT and LgBiT (Dixon et al., 2016) to SARS-CoV-2 viral protein antigens (**Figure 1A**). Since an antibody has two Fragment Antigen Binding (Fab) arms, incubating serum with 1:1 mixed SmBiT and LgBiT biosensors will result in half of the anti-viral antibodies binding LgBiT with one Fab arm, and SmBiT with the other Fab arm. This hetero-bivalent interaction localizes the LgBiT and SmBiT fragments in close proximity, resulting in reconstitution of an intact, active NanoLuc enzyme for luminescence-based detection of reactive antibodies.

**Figure 1.**
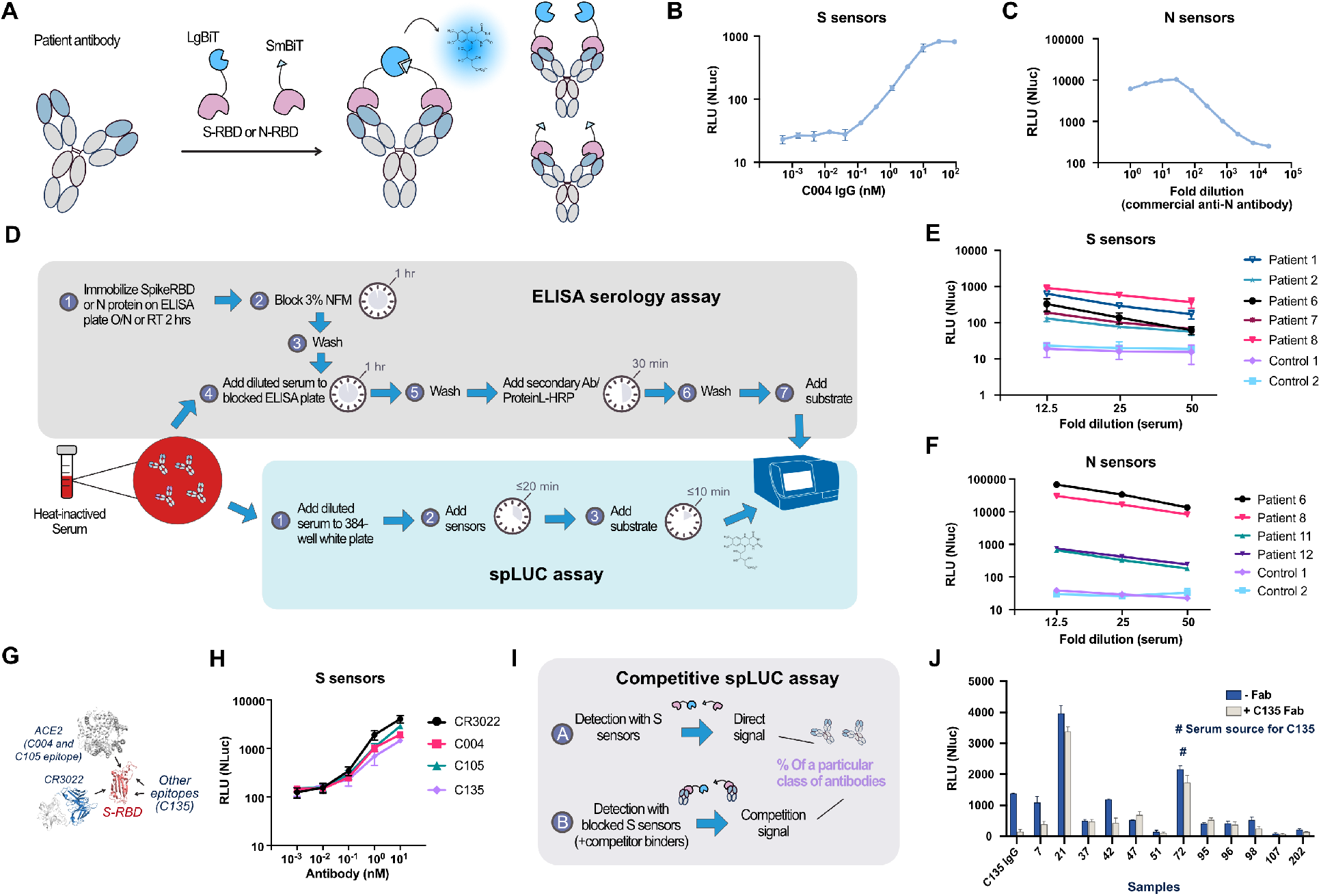
Engineering luminescent biosensors for rapid and quantitative detection of SARS-CoV-2 antibodies. **(A)** Schematic of the solution-based serology assay. Patient antibodies are incubated with SARS-CoV-2 S or N proteins fused to LgBiT/SmBiT. For the population of antibodies with one arm bound to the LgBiT sensor and the other arm bound to the SmBiT sensor, the NanoBiT luciferase enzyme is reconstituted and thus can produce active luciferase signal. **(B)** Dose-dependent spLUC signals for the recombinant anti-S-RBD antibody C004 in PBST + 10% FBS. **(C)** Dose-dependent spLUC signals for an anti-N-RBD antibody (Sino Biological, Cat#40588-T62–50) in PBST + 8% FBS. **(D)** Comparison of assay procedure between the ELISA and the spLUC assay. While the ELISA assay takes > 2 hours and involves multiple wash and incubation steps, the spLUC solution-based assay is simply completed in ≤ 30 minutes without the need for wash steps. **(E)** The S (L15+S25) sensors are able to detect antibodies in 5/5 COVID-19 recovered patients. At all dilutions tested, all 5 patients generated signal above the background signal of two control serum samples collected before the pandemic. **(F)** The N (LC+SC) sensors are able to detect antibodies in 4/4 COVID-19 recovered patients. At all dilutions of serum tested, all 4 patients generated signal above the background signal of two control serum samples collected before the pandemic. **(G)** Patient antibodies for SARSCoV-2 have various epitopes on the S-RBD (red). C004 and C105 have ACE2-competitive epitopes, while C135 and CR3022 (blue) have non-ACE2 competitive epitopes. **(H)** S sensors can detect patient antibodies of various epitopes with similar sensitivity. C004, C105, C135, and CR3022 patient antibodies were incubated with the S sensors at 10-fold antibody dilutions from 10 nM to 0.001 nM. For **(B, C, E, F, H)**, the data points represent the average of duplicates from two separate experiments. The error bars represent the standard deviation. **(I)** Schematic of antibody epitope competition assay with patient serum samples. Direct signal is compared to signal generated in the presence of the pre-incubated 1 μM Fab +1 nM sensor. **(J)** Competition assay performed with C135 Fab on twelve outpatient sera samples and recombinant C135 IgG protein. Samples were incubated with either no Fab (blue) or C135 Fab (off-white). Patient 72 (serum source of the C135 antibody) had a decrease in signal in the presence of the C135 Fab. In addition to Patient 72, patient 7, 21, 42, 98, 202 also had a decrease in signal. Bars represent the average of two replicates, error bars represent standard deviation.

We chose to develop S and N sensors for SARS-CoV-2 antibody tests because COVID-19 patient antibodies are predominantly directed against epitopes on the viral S protein, which interacts with the host receptor angiotensin-converting enzyme 2 (ACE2) and mediates viral entry (Letko et al., 2020), and the N protein, which packages the viral genome into a ribonucleocapsid (Kang et al., 2020). These two viral proteins are the primary antigens used in the current COVID-19 serological tests (Qu et al., 2020; Stadlbauer et al., 2020; Zhao et al., 2007; Byrnes et al., 2020; Amanat et al., 2020; Okba et al., 2020).

The S sensors were constructed by fusing the NanoLuc fragments to the receptor binding domain (S-RBD), which is the primary target of neutralizing antibodies (**Figure S1A, B**) (Amanat et al., 2020; Byrnes et al., 2020; Okba et al., 2020; Rosado et al., 2020). We modeled S-RBD binding to two antibodies, C105 (Robbiani et al., 2020; Barnes et al., 2020), an ACE2-competing binder, and CR3022 (Yuan et al., 2020), an ACE2 non-competing binder, to determine linker lengths (**Supplementary text, Figure S1C**). Based on the models, we constructed SmBiT fusions to S-RBD C-terminus with 15 or 25 residue Glycine/Serine (GS) linkers (S15 and S25), and LgBiT fusions to S-RBD C-terminus with 5, 15, or 25 residue GS linkers (L5, L15 and L25). These variants varied in expression yields (**Figure S1E**). Using recombinantly expressed S-RBD antibodies and ACE2 variants, we determined the optimal linker variant, enzyme concentration, buffer conditions, and impact of antibody-antigen binding affinity to signal strength (**Supplemental text and Figure S1–3**). The (L15+ S25) sensor pair at 1 nM enzyme concentration was identified as the optimal conditions for all subsequent assays.

In further characterizing the relationship between assay signal strength and antibody concentration/binding affinity, we performed ordinary differential equation modeling in R (**Supplemental text and Figure S4**). The modeling predicted a linear relationship between antibody concentration and luciferase signal (**Figure S4B**), consistent with our experimental data (**Figure 1B**). In addition, the results highlighted that the sensors at 1 nM are more sensitive to an antibody binder with a K_D_≤ 1 nM (**Figure S4B, C**). Importantly, this threshold is equivalent to the median affinity reported for polyclonal antibody repertoires (Poulsen et al., 2007; Reddy et al., 2015).

To construct the N sensors, we used the N-terminal sequence because aa 44–257 are found to be more immunogenic than the C-terminal dimerization domain (aa 258–419) (**Figure S5A**) (Zamecnik et al., 2020). In addition, dimerization promoted by the C-terminal domain may lead to high basal NanoLuc reconstitution levels. The atomic structures of N (aa 44–180) (Kang et al., 2020) showed the N and C termini are not in close proximity and therefore fusion at the N or C terminus may result in different sensor sensitivity (**Figure S5B**). Given this knowledge, three fusion sensor pairs were designed: (a) **LN+SN:** L/S-N(aa 44–257), (b) **LC+SC**: N(aa 44–180)-L/S, and (c) **LC2+SC2**: N(aa 44–257)-L/S, where L and S represent LgBiT/SmBiT, C represents C-terminal fusion, and N represents N-terminal fusion (**Figure S5C**). Testing on commercial polyclonal anti-N protein antibody revealed that the LC + SC and LC2 + SC2 sensors generated stronger signals over LN + SN (**Figure S5D**). The LC + SC sensors generated linear, dose-dependent signals with commercial anti-N protein antibody (**Figure 1C**).

We next designed a simple and rapid protocol to assay a pilot set of serum samples from convalescent SARS-CoV-2 patients (**Figure 1D**). Two healthy control sera collected before the emergence of SARS-CoV-2 virus were also tested. Serial dilutions (1:12.5, 1:25, and 1:50) of heat-inactivated sera were measured using S or N sensors. Robust, dose-dependent luminescence signal was observed across all serum concentrations tested, with the 12.5-fold dilution showing the highest signal (**Figure 1E, F**). The S (L15+S25) sensors generated signal for all five patients tested (**Figure 1E**). The N (LC+SC) sensors detected patient antibodies from all four patients tested (**Figure 1F**). However, the N (LN+SN) sensors only detected antibodies from two patient sera samples that had the strongest seropositivity (**Figure S5E**), which further confirmed a C-terminal fusion enhances NanoLuc reconstitution relative to the N-terminal fusion.

### Competitive spLUC assay to profile epitope-classes of antibodies

In addition to a test to determine total binding antibodies, an assay that allows profiling of epitope classes of antibodies can be highly valuable. In this regard, competitive ELISA assays developed by us and others have enabled characterization of percentage of ACE2-competitive antibodies (Byrnes et al., 2020; Tan et al., 2020a). These assays can potentially serve as surrogate viral neutralization tests. However, S-RBD is known to have multiple additional neutralization epitopes outside of the ACE2-binding site. An assay that allows for rapid, unbiased profiling of those alternative epitopes could unveil further details of a patient’s humoral response to neutralize SARS-CoV-2.

We first show that spLUC assay can detect antibodies binding to various epitopes on S-RBD (**Figure 1G**). We expressed and tested four reported neutralizing antibodies which bind to three distinct epitopes on S-RBD. This includes: C004 and C105 (Robbiani et al., 2020), which are ACE2-competitive binders; CR3022 (Yuan et al., 2020), which binds at a cryptic site outside of the ACE2-binding site; and C135 (Robbiani et al., 2020), which does not compete with C004, C105, CR3022 or ACE2-Fc, representing a unique binding epitope on S-RBD (**Figure S6**). All four IgG antibodies generated dose-dependent luminescence signals at ³ 0.1 nM concentrations (**Figure 1H**).

We then designed a competitive spLUC assay to determine presence of a specific epitope class of antibodies (**Figure 1I**). Out of the four antibodies tested, C135 represents an unconventional and less understood epitope class. It neutralizes very potently (IC50 = 17 ng/ml) and could be potentially used as in combination with other ACE2-competitive binders as a cocktail therapy. We converted C135 IgG to a single binding arm Fab binder, and pre-incubated 1 μM of C135 Fab with the S sensors to generate “blocked sensors”. By comparing signal between the original and the “epitope masked” sensors, we can determine how much signal from a patient’s sample corresponding to antibodies with a similar epitope (**Figure 1I**). We assayed 12 patient serum samples with representative high, medium, and low anti-S-RBD antibody levels at a 1:25 dilution of serum. IgG C135 served as a control for competition with Fab C135. Indeed, the luminescence signal of IgG C135 was reduced by ∼90% with the blocked sensors, which provided a validation of this method. Sera 7, 21, 42, 72, 98 and 202 showed a decrease in luminescence signal, indicating they likely have C135-competitive antibodies (**Figure 1J**). Patient #72 was the source for identifying C135 (Robbiani et al., 2020) and indeed showed reduction in the spLUC signal when competed with Fab. These results suggested that antibodies recognizing this unconventional, neutralizing S-RBD epitope are present in a significant proportion of patient samples. Performing this competitive serology assay with different competitive Fab antibodies in an expanded patient cohort could further our understanding of the distribution of epitopes on S-RBD as well as the correlation between binding epitopes and clinical outcomes.

### Characterization of larger cohorts of serum/plasma samples using the spLUC assay

We next applied this new assay in an expanded number of patients (**Figure 2**). First, to determine assay cutoff values and specificity, which reflects how well an assay performs in a group of disease-negative individuals, we performed the tests on three cohorts of negative control samples (Total n = 144), which include mainly healthy individual samples, 12 seasonal coronavirus patient samples, and 20 flu vaccine pre- and post-vaccination samples. All controls were collected before the SARS-CoV-2 pandemic. These controls generated significantly lower luminescent signals than the COVID-19 patient sera samples (**Figure 2A, B**). The range, median, mean and standard deviation values were calculated, and stringent cutoff values were determined by calculating the mean plus three standard deviations (**Table S1**). With these determined cutoffs, we calculated the specificity of the S sensors (1:12.5 serum dilution) to be 100% (56/56), and the N sensors (1:12.5 serum dilution) to be 99.2% (119/120).

**Figure 2.**
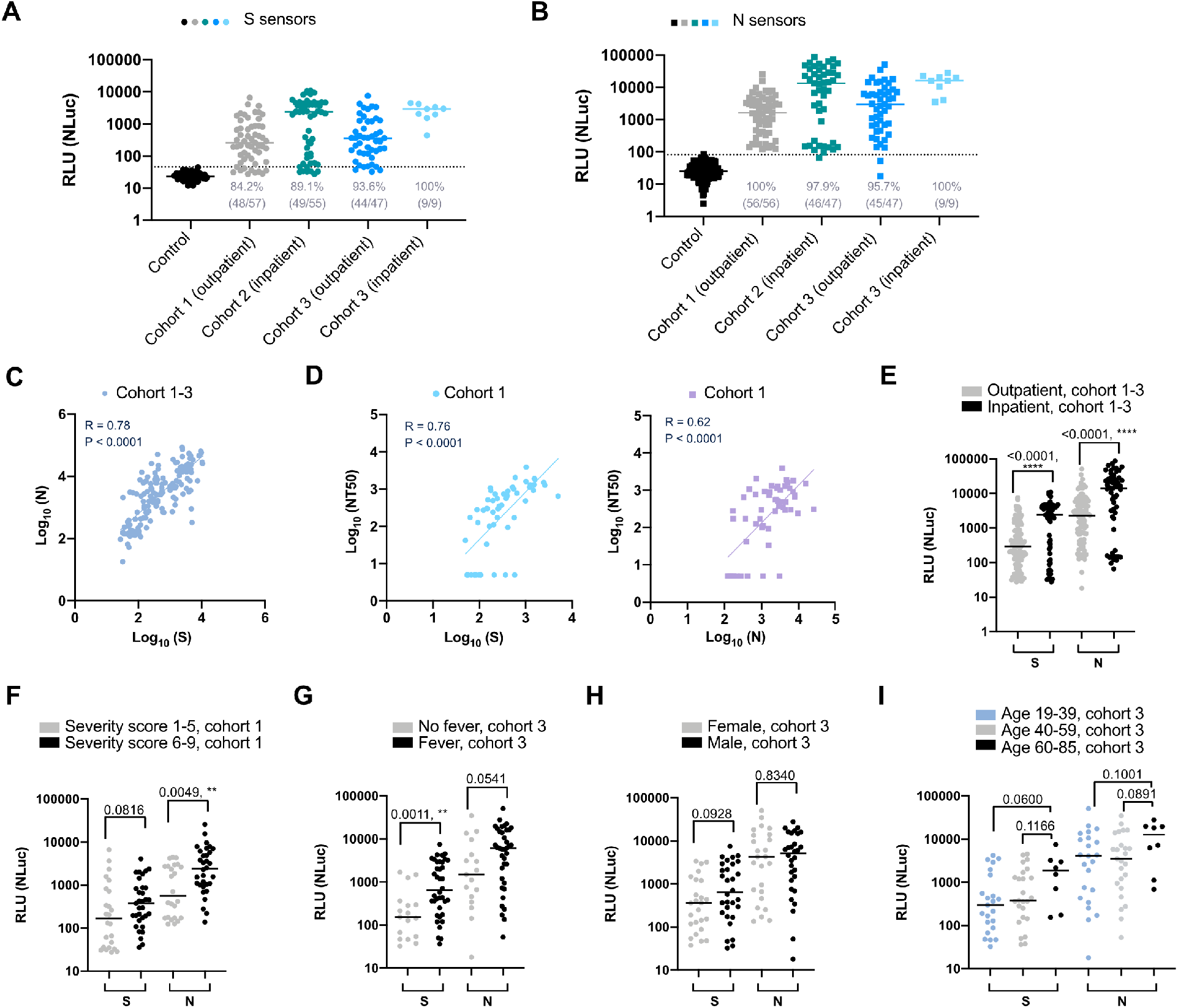
Characterization of outpatient and inpatient serum samples using the spLUC test. Cohort 1: samples drawn during the convalescent phase of an outpatient group, Cohort 2: samples drawn during the acute phase or the convalescent phase of a hospitalized group, and Cohort 3: samples drawn during the convalescent phase of a mixed inpatient and outpatient group. A 10-base logarithmic scale conversion was applied to all the solution assay signals for the correlation analysis unless otherwise specified. **(A)** SpLUC assay tested on expanded COVID-19 patient cohorts with S sensors at 1:12.5 serum dilution. Dots represent the average between two technical duplicates. Lines represent median values. The inpatient samples showed significantly higher antibody titers than the outpatient cohorts. **(B)** SpLUC assay tested on expanded COVID-19 patient cohorts with N sensors at 1:12.5 serum dilution. The inpatient samples showed significantly higher antibody titers than the outpatient cohorts. **(C)** A positive correlation (R = 0.78) was observed between S sensor signal and N sensor signal in the three cohort samples. All cohorts individually presented a similar trend (**Figure S8**). Line represent linear regression. **(D)** Correlation of spLUC signals (cohort 1) to neutralization efficiency (Robbiani et al., 2020). S sensor signal (blue) and N sensor signal (purple) is plotted against 50% maximal neutralization titer (NT50). Both show positive correlation (R = 0.76 for S and NT50 and R = 0.62 for N and NT50). **(E)** Inpatients show significantly higher signal over outpatients in all three cohorts (p < 0.0001). **(F)** Patients from cohort 1 that reported higher disease severity (6–10 vs 1–5) had higher antibody titer for both S and N sensors and the difference for N sensors is statistically significant (p = 0.0049). g, Higher overall antibodies titers were observed in patients that reported fever compared to no fever patients for cohort 3. Lines represent median values. This difference was statistically significant for the S sensors (p = 0.0011) but not N sensors. **(H)** Slightly higher overall antibodies titers were observed in females compared to males for cohort 3, although the differences were not statistically significant. There is a similar trend for cohort 1 (**Figure S9A**). The difference was more obvious for S sensors. Lines represent median values. **(I)** For cohort 3, there is a slightly higher level of antibodies in the 60–85 age group compared to 19–39 and 40–59. There is a similar trend for cohort 1 (**Figure S9B**). The differences were not statistically significant. Lines represent median values. For A, B and F-I, the Mann-Whitney test P values for each comparison are labeled on top of the datasets. For c-d, the Spearman R values and P values are labeled in the graphs. For all figures, dots represent the average of two technical replicates. Horizonal lines represent median values. For cd, lines represent linear regression.

We then used the spLUC assay to study three additional cohorts of patient samples (**Figure 2A, B**). Cohort 1 is an outpatient cohort recruited at the Rockefeller University Hospital (Robbiani et al., 2020). The samples were collected from individuals free of COVID-19 symptoms for ³14 days. The S sensors showed 84.2% (48/57) sensitivity, and the N sensors showed 100% (56/56) sensitivity. Cohort 2 samples are consisted of remnant sera from COVID-19 patients within Kaiser Permanente Hospitals of Northern California. These samples were drawn in any phase of infection, including the early acute phase. A subset of these patients, who may have not fully seroconverted at the time of sampling, had lower S sensor or N sensor signals compared to others in the spLUC assays. The sensitivities of the assays were 89% (49/55) for S sensors and 98% (46/47) for N sensors. Cohort 3 patients were part of the LIINC (Long-term Impact of Infection with Novel Coronavirus) study from San Francisco General Hospital and included plasma of a mixture of outpatient and inpatient samples drawn in the convalescent phase of the disease. With the S sensors, we detected antibodies in 94% (44/47) of outpatient samples and 100% (9/9) of inpatient samples. With the N sensors, we detected antibodies in 96% (45/47) of outpatient samples and 100% (9/9) of inpatient samples. For all cohorts, the S and N signals show a strong correlation (**Figure 2C, Figure S8**). Consistent with previous findings, we observed varying degrees of anti-S and N antibody seropositivity between patients (**Figure 2A, B**), which reflects a wide range of patient humoral response to this virus (Long et al., 2020; Lynch et al., 2020).

Importantly, we observed strong correlation of spLUC assay results to anti-Fab and anti-IgG SRBD ELISA signals (**Figure S7A-C**, R = 0.43–0.91). A base-10 logarithmic scale conversion was applied to the spLUC assay signals for the correlation analysis to ELISA signals. This non-linear correlation between the spLUC and ELISA assays is likely due to signal compression in ELISAs at high antibody concentrations (Abcam, ELISA guide). For all cohorts, the S sensor seronegative samples also had very low signals in S-RBD ELISA assays (**Figure S7D–F**), which confirmed the presence of low levels of anti-S-RBD antibodies in these sub-cohorts of patients. Interestingly, the correlations to IgM signals were much weaker (**Figure S7A–C**). It is possible that IgM was not sensitively detected by the spLUC assay due to the weaker affinities of the individual binding arms in IgMs (Mäkelä et al., 1970), or that the IgG response dominated the signal in many of the tested patients.

One of the key uses of a highly sensitive serology assay is to grade the quality of convalescent sera to neutralize virus (Krammer and Simon, 2020). In cohort 1, our analysis showed the S sensor signals correlated with the half-maximal neutralizing titers (NT50s) reported by Robbiani et al(**Figure 2D, left panel**), which is consistent with previous studies on the relationship between anti-S antibody titers and neutralization potency (Seow et al., 2020; Wajnberg et al., 2020; Amanat et al., 2020; Robbiani et al., 2020). Interestingly, we found that the N sensor signals showed a similar correlation with NT50 (**Figure 2D, right panel**). Our results indicate determining either anti-S or anti-N seropositivity is a general means to assess the neutralization potential of sera samples.

To try and gain clinical insights from our results, we analyzed our spLUC data in the context of clinical and demographic features. First, the degree of seropositivity for inpatient samples was significantly higher than that of outpatient samples (**Figure 2A, B, E**). Disease severity scores and fever were also associated with a stronger antibody response (**Figure 2F and G**). These results indicated a direct correlation of disease severity and adaptive immune response consistent with previous studies (Zhao et al., 2020; Robbiani et al., 2020; Cervia et al., 2020; Lynch et al., 2020; Long et al., 2020; Seow et al., 2020; Klein et al., 2020). In addition, males had slightly higher antibody titer than females in both cohort 1 and 3 especially for anti-S antibodies, although the differences were not statistically significant (**Figure 2H, Figure S9A**). This finding was consistent with studies by Klein et al (Klein et al., 2020) and Robbiani et al (Robbiani et al., 2020), but different from Zeng et al (Zeng et al., 2020), which reported females with severe disease developed more antibodies than men with severe disease. This difference might be due to differing selection criteria of patient cohorts. Lastly, patients of age 60–85 showed a higher trend of antibody response compared to those in the 19–39 and 40–59 age brackets, but the difference was not statistically significant (**Figure 2I, Figure S9B**). Similar findings on the impact of age have been reported previously (Whitman et al., 2020; Lassaunière et al., 2020). These results highlight that demographic and clinical features affect the antibody response of COVID-19 patients. A longer-term, systematic, and population-level serological analysis is needed to further illuminate the variables that affect patient humoral response to SARS-CoV-2.

Collectively, our assay showed high sensitivity and specificity for all three representative cohorts of serum/plasma samples (inpatient, outpatient, acute phase, convalescent phase), with an overall specificity of 100% (S sensor) and 99% (N sensor), and sensitivity of 89% (S sensor) and 98% (N sensor). These values are comparable or superior to reported values for laboratory ELISA and lateral flow tests (Whitman et al., 2020; Lassaunière et al., 2020).

### Adapting the assay for low-resource settings and expanded sample types

Lastly, we adapted our assays to begin to meet the clinical needs in remote and low-resources settings and for point-of-care or large-scale deployment. While the current properties of the assay meet most of the requirements for deployment in these types of settings, we tested to see if the reaction time (30 minutes), reagent format (frozen aliquots of sensors), and sample type (serum/plasma) could be further optimized.

We first tested if our initial reaction times (20-minute sensor/antibody incubation and 10-minute incubation with substrate, **Figure 1D**) are necessary and optimal. CR3022 (10 nM) was incubated with 1 nM S sensors for 5, 10, 15, and 20 min, followed by luciferase substrate addition and incubation for 0, 2, 4, 6, 8, and 10 minutes (**Figure 3A**). All time points resulted in bright luminescence signal, suggesting that the assay could be completed in as short as 5 minutes.

**Figure 3.**
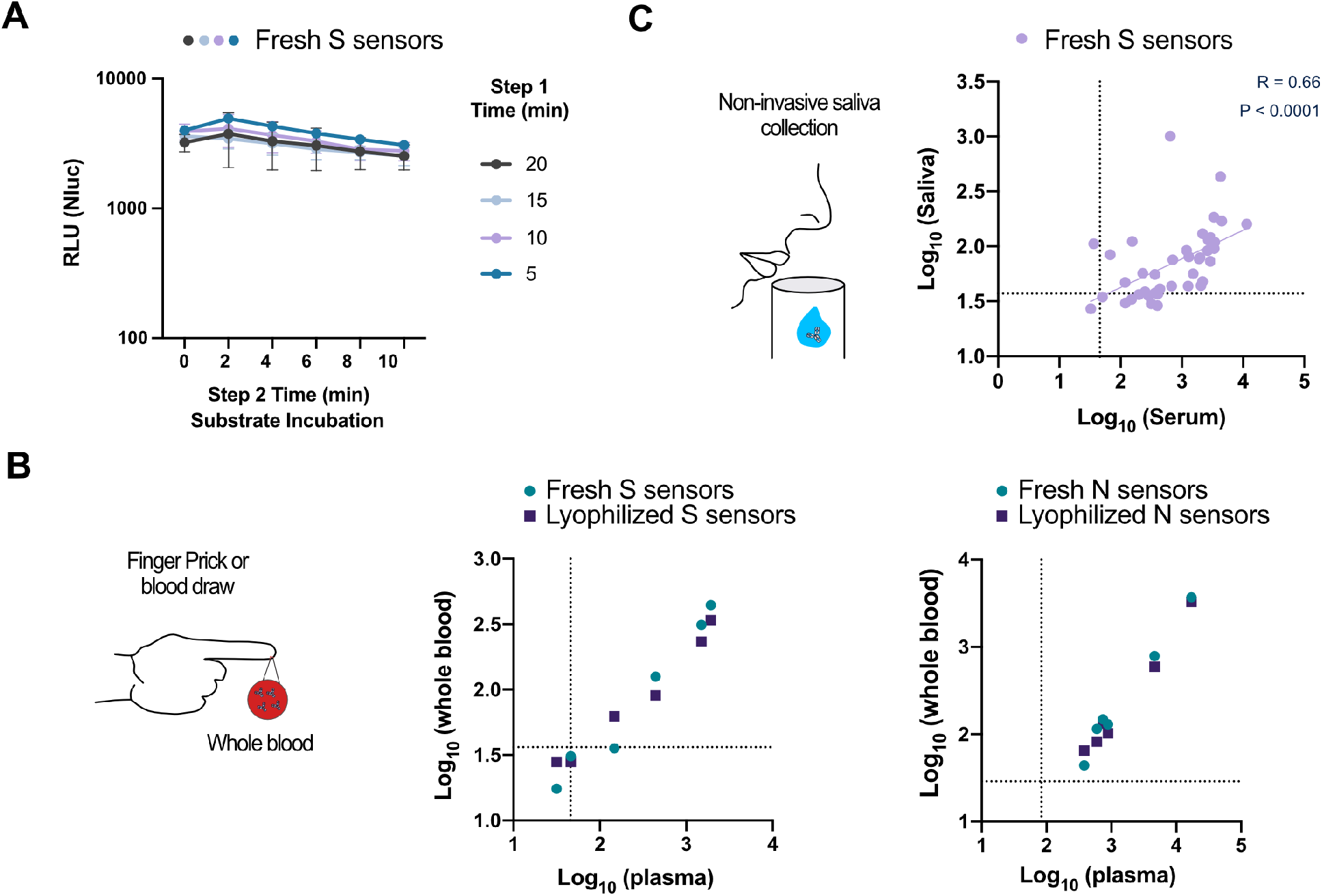
Adapting the assay for whole blood and saliva sample types. (A)spLUC assays can be accomplished in as short as 5 minutes. CR3022 (10 nM) was incubated with S sensors for 5, 10, 15, or 20 min. Luciferase substrates were then added and incubated with the reaction mix for 0, 2, 4, 6, 8 or 10 min. All reactions showed bright luminescence signal. Error bars represent the standard deviation. **(B)** The spLUC assay is compatible with whole blood samples and show similar signal in the corresponding plasma samples with both fresh and lyophilized sensors (R = 0.94 for S sensors, R = 1 and 0.98 for N sensor fresh and lyophilized sensors, respectively). **(C)** Anti-S antibodies were detected in saliva samples with moderate sensitivity (33/42, 79%). The signals from saliva samples positively correlated with corresponding serum samples (R = 0.66, p< 0.0001). For a-c, each dot represents the average of two technical replicates.

We then tested if the sensors can be lyophilized for ambient temperature storage and transportation. Although a small quantity (0–30%) of S sensors and N sensors were lost due to the lyophilization process (**Figure S10A**), both the lyophilized S and the N sensors can still robustly detect recombinant IgG or patient antibodies in serum with similar sensitivities seen for the fresh sensors (**Figure S10B, C**).

Finally, we sought to determine if the spLUC assay could be compatible with other sample types. First, whole blood samples were collected from six convalescent COVID-19 patients and plasma samples were prepared in parallel for comparison (**Figure 3B**). Remarkably, although the overall signals were lower from whole blood samples, all six samples generated N sensor signals and four had S sensor signals above control levels with the lyophilized sensors (**Figure 3A**). In comparison, all six patients generated N sensor signals and five had S sensor signals above cutoff values from the plasma samples. Strong correlations were observed between the whole blood signals and the plasma signals (R > 0.9). Fresh and lyophilized sensors showed very little difference in performance.

Next, we tested the potential of using saliva as an input. To determine conditions, we added varying concentrations of the CR3022 antibody into saliva from a healthy individual (**Figure S11**). We saw a significant reduction in sensitivity for undiluted saliva relative to buffer alone, but remarkably no loss in sensitivity when the saliva was diluted 1:2 in PBS buffer. We then tested 42 saliva samples at 1:2 dilution with the S sensors. We increased the reaction volume from 20 to 100 μl and the luminescence signal integration time from 1000 ms to 5000 ms for better sensitivity, as lower antibody concentrations are expected from saliva samples (Randad et al., 2020). Out of the 42 samples, 33 had signals above the two healthy saliva controls, indicating a 79% assay sensitivity (**Figure 3C**). A moderate correlation of saliva signal with corresponding serum signals was observed (R = 0.66), consistent with recent reports (Faustini et al., 2020). These results highlight the potential of using lyophilized sensors and whole blood or saliva samples as a convenient diagnostic workflow for rapid and quantitative point-of-care antibody testing amenable to broad population deployment or applications in resource-limited areas.

### DISCUSSION

As the SARS-CoV-2 virus continues to spread, the need will continue to grow for serology assays to determine not only the scope of infection, but also vaccine efficacy during clinical trials and after large-scale vaccine deployment. We present here spLUC, a simple (no wash, two-step of reagent addition), sensitive (≥98%), specific (≥99%), fast (as short as 5 minutes), low-input sample volume (1 μl per reaction), low-cost (∼15¢ per reaction), and quantitative solution-phase serological assay to detect antibodies against S and N proteins. We were able to test 159 patient samples across three different cohorts with varying clinical and demographic features. Our results enabled association analysis between these features (e.g. hospitalization, disease severity, presence of fever, gender, age), demonstrating the promise of this rapid assay to generate large datasets to better understand factors that modulate the humoral response following SARS-CoV-2 infection.

The quantitative and solution-based nature of the spLUC assay allows convenient assay variations. We presented a competitive spLUC assay using epitope masked S sensors and used it to study the prevalence of an unconventional neutralization epitope in the S-RBD domain. This competitive spLUC assay has the potential to serve as a surrogate virus neutralization assay and to unveil details of the interaction of patient antibodies to viral antigens.

Robust ELISA-based assays such as the one developed by Krammer and co-workers have enabled tremendous progress of COVID-19 serological studies (Amanat et al., 2020; Stadlbauer et al., 2020), but these assays are still laborious with multiple wash steps, which limits their feasibility for population-scale sero-surveillance, point-of-care diagnostics, and deployment in countries or remote areas that have limited access to analytical equipment and reagents. The spLUC assays have important features amenable to all these applications. We have shown that our reagents are not only compatible with lyophilization for easy transport and storage, but can also readily detect antibodies directly from whole blood samples and saliva samples. With simple pipettes and a battery-supported portable luminometer (e.g. 32526–11 Junior LB9509, Berthold Technologies), the spLUC assay could be readily established at care centers or in the field worldwide, regardless of infrastructure. To this end, we are currently collaborating with bioengineers to develop portable luminometers that can be manufactured at low cost but provide equal or better detection sensitivity.

Another important strength of our approach is the modularity. We expect, with modifications to the sensor designs, that our strategy can be readily adapted to develop rapid serological tests for immunity against virtually any infectious disease that elicits an antibody response for which the protein antigen is known. Future development of our spLUC assay includes exploring orthogonal split enzyme systems to allow multiplexing of assays. For instance, split ß-lactamase, used by Huang and co-workers for detecting herpes simplex virus antibodies (Fry et al., 2008), can provide an orthogonal readout to luminescence. We envision that such multiplexed assays could be used to develop broad-spectrum serological assays to simultaneously detect immunity against multiple infectious diseases.

In summary, we have taken a structure-based protein engineering approach to design novel split enzyme-fused sensors. These biosensors enable spLUC, a next-generation SARS-CoV-2 antibody test suited for population-scale sero-surveillance, epitope mapping of patient antibody responses, and testing in resource-limited areas. Future efforts will focus on continued evaluation of alternative sample sources and development of similar split enzyme-based serological approaches for a range of infectious diseases.

## Data Availability

All data described in the manuscript is available upon request.

## ACKNOWLEDGEMENTS

We acknowledge the members of the Wells Lab, especially those involved in our COVID-19 research program. We also acknowledge Dr. Michel Nussenzweig, Dr. Marina Caskey, and Dr. Christian Gaebler (Rockefeller University), as well as Dr. Colin Zamecnik and Dr. Joseph DeRisi (University of California, San Francisco) for providing the first cohort of convalescent sera. We thank Dr. Michel Nussenzweig and Dr. Davide Robbiani for providing plasmids for C004, C105, and C135 IgGs. We additionally thank Dr. Peter Kim for providing the plasmid for CR3022. We thank Patrick Wilson (U. Chicago) for the seasonal coronavirus control samples. We thank Dr. Michael Lin (Stanford University) for helpful discussions regarding split enzyme systems. We also thank Charles Chiu (UCSF) for helpful discussions. We thank all the patients for their participation in this study.

J.A.W. is grateful for funding from the Harry and Dianna Hind Endowed Professorship in Pharmaceutical Sciences and the Chan Zuckerberg Biohub that helped support this work. The National Science Foundation Graduate Research Fellowship Program supported S.K.E (1650113) and I.L (2017244707). Postdoctoral Fellowship support included a National Institutes of Health National Cancer Institute F32 (5F32CA239417 to J.R.B.), a Merck Fellow of the Damon Runyon Cancer Research Foundation (DRG-2297–17 to X.X.Z.), a Merck Postdoctoral Research Fellow from the Helen Hay Whitney Foundation (S.A.L.), and a National Institutes of Health K99/R00 (1K99GM135529 to A.A.G.). The LIINC cohort study was funded by NIH/NIAID 3R01AI141003–03S1 (T.J.H.). T.T.W was supported in part by Fast Grants, CEND COVID Catalyst Fund, the NIH/NIAID (U19AI111825 and R01AI139119) and by the Rockefeller University Center for Clinical and Translational Science Grant # UL1 TR001866. We would like to acknowledge funding from the Chan Zuckerberg Biohub, Rapid Response as well.

## AUTHOR CONTRIBUTIONS

S.K.E and X.X.Z conceived the study and designed the experiments. S.K.E., X.X.Z., and J.R.B. analyzed data and wrote the manuscript. S.K.E. performed the experiments unless otherwise stated.

X.X.Z. performed structure modeling. J.R.B. performed the anti-Fab ELISA experiments and provided advice for the whole blood work. A.J.M. performed the in silico differential equation modeling. I.L., K.P, and S.A.L. helped with expression, purification, and performed the ACE2 epitope binning experiment. J.E.G. and A.A.G. designed, expressed, and purified the higher affinity ACE2 mutant. T.T.W. provided patient sera and control sera samples. T.J.H., M.J.P., B.G., N.I., L.T., and K.T. provided patient samples, oversaw LIINC samples collections, sample processing, sample maintenance, and cohort design. B.G., C.M.T. and K.K.L. provided helpful discussions. J.A.W. supervised the research. All authors provided edits and approval of the final manuscript version.

## METHODS

All data described in the manuscript is available upon request.

### Plasmid construction

Plasmids were constructed by standard molecular biology methods. The DNA fragments of Spike-RBD, N protein, ACE2, and LgBiT were synthesized by IDT Technologies. The SmBiT tag was generated by overlap-extension PCR. The Spike-RBD-5/15/25aa-LgBiT-12xHisTag, Spike-RBD-15/25aa-SmBiT-12xHisTag, N protein(44–180)-10aa-LgBiT-12xHisTag, N protein(44–180)-10aa-SmBiT-12xHisTag, LgBiT-10aa-N protein(44–257)-12xHisTag, and SmBiT-10aa-N protein(44–257)-12xHisTag were generated by subcloning into a pFUSE-12xHisTag vector (adapted from the pFUSE-hIgG1-Fc vector from InvivoGen). The ACE2-Fc fusion plasmids were generated by subcloning the gene fragments of ACE2 and mutant into the pFUSE-hIgG1-Fc vector. The C004, C105, and C135 IgGs LC and HC plasmids were a generous gift from the Nussenzweig lab (Rockefeller University). The CR3022 IgG plasmids were a generous gift from the Kim lab (Stanford) and the Wilson lab (Scripps). The C135 Fab was cloned by removing the Fc domain from the HC plasmid. Complete plasmid sequences are available upon request.

### Expression and protein purification

All proteins were expressed and purified from Expi293 BirA cells according to established protocol from the manufacturer (Thermo Fisher Scientific). Briefly, 30 μg of pFUSE (InvivoGen) vector encoding the protein of interest was transiently transfected into 75 million Expi293 BirA cells using the Expifectamine kit (Thermo Fischer Scientific). For the IgG and Fab proteins, 15 μg of each chain was transfected. Enhancer was added 20 h after transfection. Cells were incubated for a total of 3 d at 37 °C in an 8% CO_2_ environment before the supernatants were harvested by centrifugation. Fc-fusion proteins were purified by Protein A affinity chromatography and His-tagged proteins were purified by Ni-NTA affinity chromatography. Purity and integrity were assessed by SDS/PAGE. Purified protein was buffer exchanged into PBS and stored at −80 °C in aliquots.

### Solution serology protocol for in vitro, serum, blood, and saliva samples

LgBiT and SmBiT sensors for either the Spike or N protein were prepared at a final concentration of each sensor at 2nM in PBS + 0.05% Tween-20 + 0.2% BSA (PBSTB). For in vitro IgGs or ACE2-Fc, the samples were prepared at 1:10 dilutions in PBSTB unless otherwise specified. Serum and blood samples were diluted to 1:12.5 for both the S and N sensor samples in PBSTB unless otherwise specified. Healthy individual saliva was spiked in with CR3022 and used undiluted or diluted 1:2 in PBSTB. 10 μL of the 2 nM sensor mix and 10 μL of the sample were combined in a 384 Lumitrac white plate (Greiner), skipping every other well and row to avoid potential bleedover in signal. The plate was mixed on a plate shaker for 20 minutes. NanoLuc substrate was diluted according to protocol 1:50 in NanoLuc dilution buffer (Promega) and 15 μL was added to each well, followed by a 10-minute incubation period for the signal to stabilize. Luminescence was measured on a Tecan M200 infinite plate reader with an integration time of 1000 ms.

### Competition serology protocol for in vitro and serum samples

The competition serology assay was performed similarly to the solution serology assay except that the S sensors were individually preincubated at 4 nM with 4 μM of either C004 Fab, C105 Fab, or C135 Fab for the in vitro competition assay and C135 Fab only for the serum competition assay. The two sensors + Fab were combined 1:1 to make a 2 nM mix, and 10 μL of this mix was added to the assay as described above.

### Epitope binning experiment

Biolayer interferometry data was measured using an Octet RED384 (ForteBio). Biotinylated Spike RBD protein was immobilized on the streptavidin (SA) biosensor (ForteBio). After blocking with biotin, the sensor was loaded with one IgG followed by another IgG or ACE2-Fc to determine epitope binning. PBS with 0.05% Tween-20 and 0.2% BSA was used for all diluents and buffers.

### Spike protein ELISA assay

The Spike ELISA assay was performed as previously described. Briefly, 384 Maxisorp plates were coated with 100 μL of 0.5 μg/mL Neutravidin for 1 hr. The plate was washed 3 times with PBS + 0.05% Tween-20 (PBST) followed by incubation with 20nM S-RBD for 30 minutes. Following 3 washes, the plate was blocked with 3% non-fat milk in PBS for 1 hour. The plate was washed 3 times before the addition of 1:50 dilutions of serum in 1% non-fat milk for 1 hour. After 3 washes, secondary anti-Fab, anti-IgG, or anti-IgM antibody was added and incubated for 30 minutes before the addition of TMB for 3 minutes. The reaction was quenched with 1 M phosphoric acid and absorbance was read on a Tecan M200 infinite plate reader at 450 nm.

### Lyophilization of sensors

The S and N protein sensors were flash frozen in liquid nitrogen at concentrations between 10–60 μM in 10 μL. A small hole was poked into the caps of the samples and left on a Benchtop K (VirTis) lyophilizer overnight. The next day the sensors were reconstituted in 10 μL of ddH_2_O and concentration was verified by nanodrop.

### Serum, plasma, whole blood, and saliva samples

The initial small patient cohort was a generous gift from the Wilson lab (UCSF) and heat inactivated at 56°C for 1 hour before storage at –80°C. The first (outpatient) sample serum set (cohort 1) was a generous gift for the Wilson lab (UCSF) and Nussenzweig lab (Rockefeller). These samples were heat inactivated at 56°C for 1 hour and stored at 4°C in a 1:1 dilution in 40% glycerol, 40 mM HEPES (pH 7.3), 0.04% NaN_3_, in PBS. The second (inpatient) sample serum set (cohort 2) was a generous gift from the T. Wang lab (Stanford) and were stored at –80°C as pure serum samples. The third plasma cohort (cohort 3) and blood samples were generous gifts from the Greenhouse lab (UCSF) and Henrich Lab (UCSF) as part of the LIINC study. The plasma samples were stored at 4°C in a 1:1 dilution in 40% glycerol, 40 mM HEPES (pH 7.3), 0.04% NaN_3_, in PBS. The whole blood was stored undiluted at 4°C. Healthy blood samples were purchased from Vitalent and stored undiluted at 4°C. The saliva samples were obtained unstimulated, unexpectorated saliva and were stored at –80°C. Before assayed, the samples were thawed and centrifuged at 9,000g to remove any insoluble or coagulated matter. Control saliva from Nov 2019 was purchased from Lee Biosciences, stored at –20°C, and processed similarly.

### Study Approval of Patient Samples

All patient samples were obtained using protocols approved by the UCSF, Stanford University, and Rockefeller University Institutional Review Boards and in accordance with the Declaration of Helsinki. Samples were de-identified prior to delivery to the lab where all assays described here were performed. Collection of remnant sera from Kaiser Permanente was approved by the Institutional Review Board of Stanford University (protocol #55718). Influenza virus vaccination samples were from a US cohort enrolled at the Rockefeller University Hospital in New York City in 2012–2013 under a protocol approved by the Institutional Review Board of Rockefeller University (protocol #TWA-0804). Samples from people with seasonal coronavirus infections were collected at the University of Chicago. Samples were de-identified serums of healthcare workers that had respiratory illnesses, were swabbed, and tested positive for common cold coronavirus infections in 2019 (U. Chicago protocol # 09–043-A).

### Data and Statistical analysis

All graphing and statistical analysis was performed in GraphPad Prism. The non-parametric Spearman correlation analysis was used in Prism to determine the correlation R value between datasets. An unpaired Mann-Whitney test was performed to determine the difference between datasets. A two-tail P value was used to determine statistical significance for all analysis. P < 0.05 was considered statistically significant.

## SUPPLEMNTAL INFORMATION

Supplemental text, Fig. S1–S11, Table S1 are attached to the end of the PDF.

## Supplementary Materials

### S sensor engineering and characterization

#### Linker modeling

We modeled S-RBD binding to two antibodies to determine the optimal linker lengths between the S-RBD domains and the SmBiT/LgBiT fusions. The antibody C105 is an ACE2-competitive binder (Figure S1C) (Robbiani et al., 2020; Barnes et al., 2020), while the antibody CR3022 does not compete with ACE2 (Figure S1D) (Yuan et al., 2020). Based on the assumption that the wing-span of antigen binding sites between Fab arms on a flexible-hinge region of an Fc are roughly ∼117–134 Å apart (Sosnick et al., 1992), and residue-to-residue distance in a linker lies between the length of tightly packed alpha-helix residues (1.5 Å) and extended beta-strand residues (3.5 Å), we estimated the total number of linker residues should be ∼30–80 amino acids. Antibodies binding to the CR3022 epitope may require a shorter linker for NanoLuc reconstitution (Figure S1D) than antibodies competitive with ACE2 (Figure S1C). Considering S-RBD has a C-terminal 15-residue loop to function as part of the linker, we constructed SmBiT fusions to S-RBD C-terminus with 15 or 25 residue Glycine/Serine (GS) linkers (S15 and S25), and LgBiT fusions to S-RBD C-terminus with 5, 15, or 25 residue GS linkers (L5, L15 and L25). These linker variants were expressed in Expi293 cells and varied in expression yields (Figure S1E). The N-terminal fusions to S-RBD were not designed because the N and C termini localize in close proximity and we hypothesized this alternative fusion design would result in similar sensor performance as the C-terminal fusions (Figure S1B).

### Optimization of enzyme concentrations, linkers and buffer conditions

We then determine the optimal enzyme concentration. A three-fold dilution series from 27 to 0.11 nM of the L15 + S25 sensors were mixed with increasing 10-fold dilutions of recombinant CR3022 (Figure S1F). After a 20-minute incubation, the NanoLuc substrate was added and allowed to develop for 10 minutes before luminescence signal was read. High sensor concentrations (27, 9, 3 nM) resulted in stronger background luminescence signal and therefore lower detection sensitivity of CR3022, due to increased basal association of the two split sensors. Meanwhile, low sensor concentrations (0.33 and 0.1 nM) generated overall less signal than 1 nM sensors because fewer sensors are captured on each antibody. As a result, sensors at 1 nM were used in all subsequent assays.

Next we queried if linker lengths affect detection sensitivity. Sensors with varied linker lengths were mixed with 10-fold dilutions of CR3022 and all resulted in dose-dependent luminescence signals (Figure S1F). Little difference in detection sensitivity was observed, except that the (L5 + S15) and (L5 + S25) linker combinations resulted in slightly decreased sensitivity at low antibody concentrations. This result indicated that we had selected a proper range of linker lengths. Based on robust signal and expression yields (Figure S1E), we chose the L15 and S25 sensor pair for subsequent assays.

Interestingly, we observed that the regular PBSTB assay buffer (PBS, 0.05% Tween-20, 0.2% m/v BSA, PBSTB) produced a higher background signal (average relative luciferase units (RLU) = 70–80) than in serum samples (RLU = 24.5). We tested if supplementing Fetal Bovine Serum (FBS) can reduce background (Figure S2). PBS + 0.05% Tween-20 (PBST) with 4–10 % FBS was found to reduce the signal (mean RLU = 21) to a level that is close to signal from 12.5% serum, and therefore can serve as a proper negative control. Both the recombinant anti-S antibody C004 and the commercial anti-N antibody (Sino biological, Cat#40588-T62–50) produced linear dose-dependent signal in this buffer (**Figure 1B and C**), which can be used to generate standard curves and calibrate the instruments for the spLUC assay.

### Impact of binding affinities

To determine whether the affinity of the target binding to S-RBD affects signal strength, we turned to two dimeric ACE2 constructs: ACE2-Fc, which is the human ACE2 peptidase domain fused to IgG1 Fc(Lui et al., 2020), and an engineered ACE2-Fc variant that binds ∼10x tighter to S-RBD (Figure S3). Overall, signal from wild-type ACE2-Fc (K_D_ = 10 nM) is weak, with signal that is more than two standard deviations above background only detected at the highest tested ACE2-Fc concentration (10 nM). Conversely, the enhanced-affinity ACE2-Fc variant (K_D_ = 1 nM) generated a dose-dependent signal from 0.1–10 nM protein concentrations and exhibited 2.6-fold higher signal observed at 10 nM relative to the wild-type ACE2-Fc. These findings indicated the sensors report the presence of not only larger quantities of anti-S-RBD binders but also higher-affinity binders. This property of the sensors suggested spLUC assay may be used to characterize binding affinities of S-RBD antibodies or ACE2 variants for therapeutic applications.

### Thermodynamic sensor model

In further characterizing the relationship between assay signal strength and antibody concentration/binding affinity, we performed ordinary differential equation modeling in R. We made assumptions such as a sensor can only be bound by one antibody, that antibody binding is non-cooperative, and that there is no detectable basal affinity of LgBiT and SmBiT at the concentrations tested (Figure S4A). The modeling predicted a linear relationship between antibody concentration and luciferase signal (Figure S4B), consistent with our experimental data (**Fig. 1B, C**).

The following set of ordinary differential equations (ODEs) was written to describe the system depicted in Figure S4A and generated the curve graphs in Figure S4B and C:

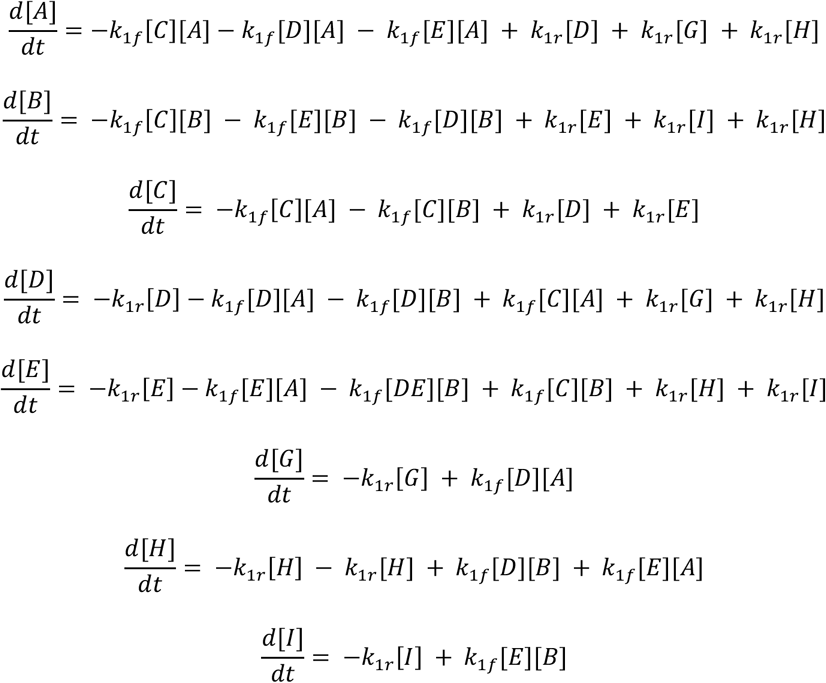

Where:

A = LgBiT sensor
B = SmBiT sensor
C = Antibody
D = Antibody/ LgBiT sensor heterodimer
E = Antibody/ SmBiT sensor heterodimer
G = Antibody/ LgBiT sensor/LgBiT sensor trimer
H = Antibody/Active Enzyme trimer (Active Enzyme)
I = Antibody/ SmBiT sensor/SmBiT sensor trimer
k_1f_ = on rate of Antibody binding to Spike
k_1r_ = off rate of Antibody binding to Spike

For simplification, we assumed the following: 1) LgBiT sensor and SmBiT sensor had no measurable interaction, 2) Antibody binding to LgBiT sensor or SmBiT sensor was non-cooperative, and 3) Antibody binding to LgBiT sensor was equivalent in rate to antibody binding to SmBiT sensor. The equations above were solved in R using the deSolve package to find the concentration of each species at equilibrium. In all cases the initial concentrations of D, E, G, H, and I were set to 0.

---

**Figure S1.**
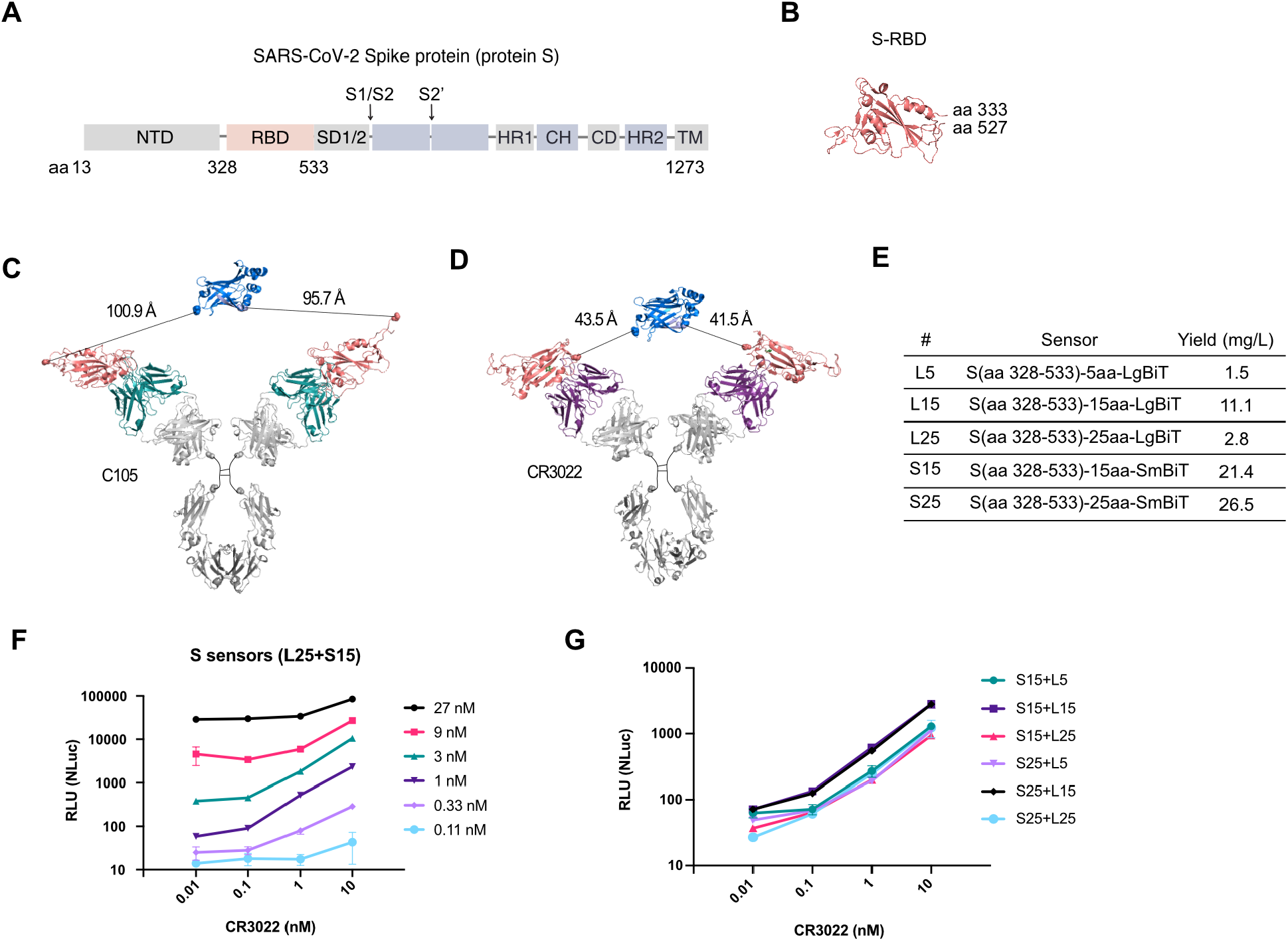
Design and characterization of S sensors. (A) Annotated depiction of the SARS-CoV-2 Spike protein. The S sensors were developed using only the S-RBD domain (aa 328 – 533, PDB: 6W41) shown in pink. **(B)** Structure of the S-RBD domain shows the N and C termini locate in close proximity. **(C, D)** Modeling of c, ACE2-competitive antibody C105 (PDB: 6XCN) binding to S-RBD-SmBiT/LgBiT sensors, and d, CR3022 (PDB: 6W41) binding to S-RBD-SmBiT/LgBiT sensors. Modeling and distance measurements were performed with PDB 6XCN, 6W41, 1N8Z, 5IBO and 5D6D in PyMOL. **(E)** Yield of the 5 Spike-NanoBiT sensor fusions. The Spike LgBiT sensors were made with 5aa, 15aa, and 25aa GS linkers (L5, L15 and L25). The Spike SmBiT sensors were made with 15aa, and 25aa GS linkers (S15 and S25). Because the N and C termini of the S-RBD domain locate in close proximity, only fusions to the C termini of S-RBD were constructed. **(F)** The S sensors are most sensitive at 1 nM for detecting CR3022 in solution compared to higher or lower sensor concentrations. **(G)** S sensors with varied linker lengths resulted in very similar signal strength in detecting CR3022.

**Figure S2.**
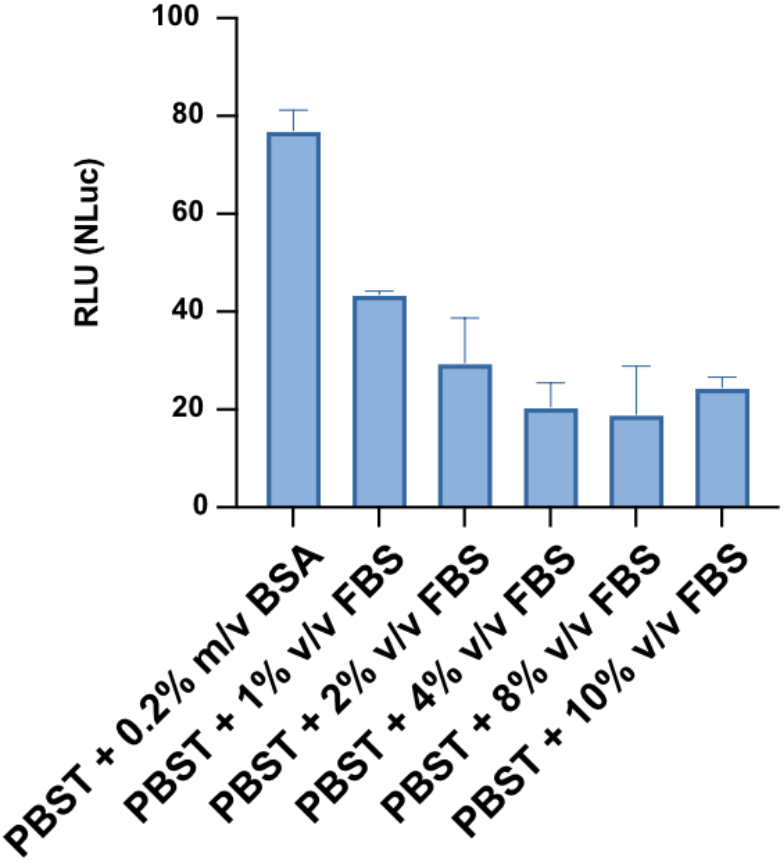
Supplementing FBS reduces background signal in spLUC assays. PBST with 4–10% FBS can be used as a negative control for serum samples as it shows similar signal suppression.

**Figure S3.**
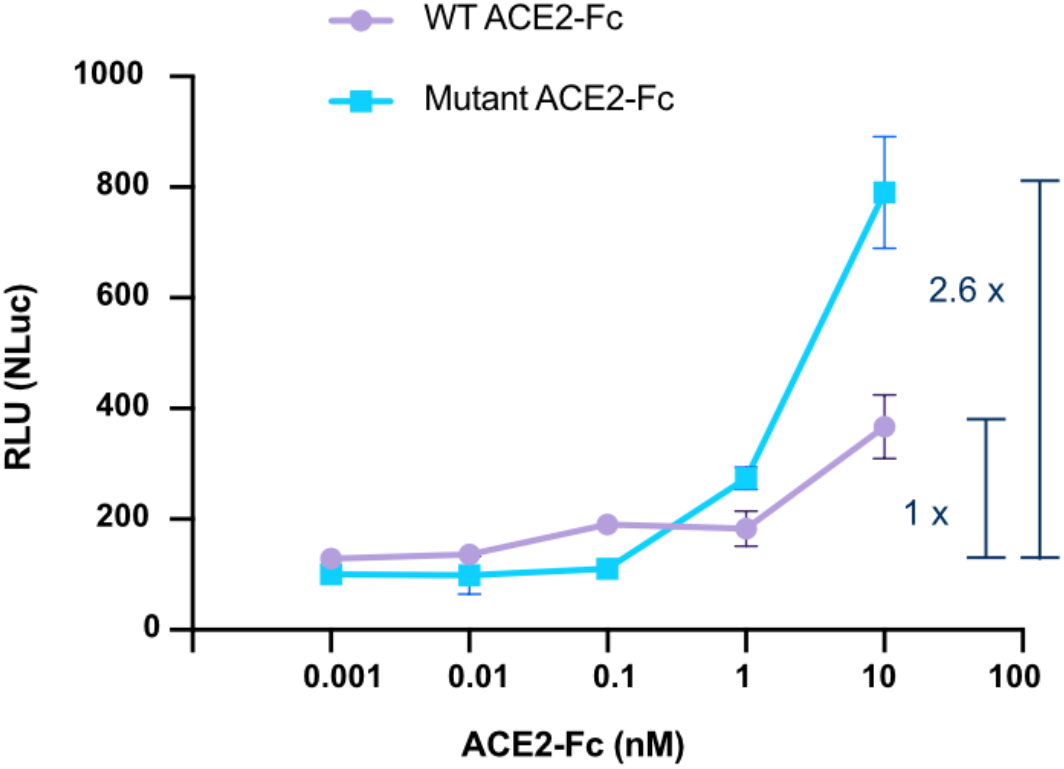
The biosensors are more sensitive to high-affinity binders. The ACE2-Fc variant which bind 10-fold tighter to S-RBD generated ∼3-fold higher signal at 10 nM protein concentration comparing to WT ACE2-Fc.

**Figure S4.**
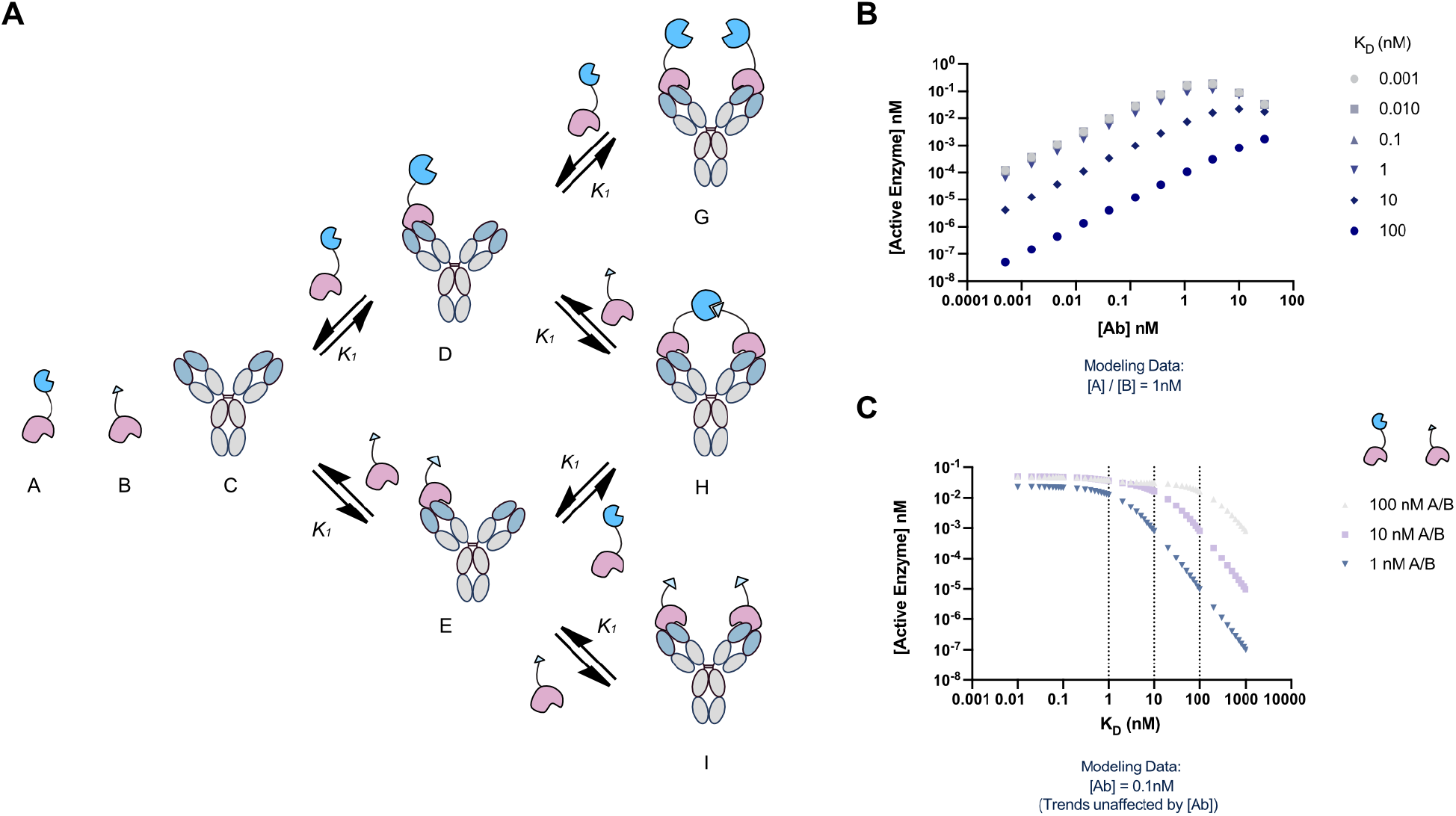
ODE models predict a linear, dose-dependent response and KD dependence of the luminescence signal. **(A)** Antibody (C) and sensor components (A and B) are in thermodynamic equilibrium with enzymatically inactive (D, E, G, and I) and active (H) sensor bound species.**(B)**At 1 nM starting concentration of sensor ([A] and [B]), spLUC assays are predicted to generate signals linearly correlated to a broad range of antibody concentrations ([Ab]). Signal is predicted to be insensitive to antibody concentrations for antibodies with high affinity for the sensor (≤ 1nM), but weaker affinity antibodies (K_D_ > 1 nM) will result in significantly lower levels of reconstituted enzyme. **(C)** At K_D_ values equivalent or higher than the sensor concentrations, the spLUC signals are predicted to drop significantly.

**Figure S5.**
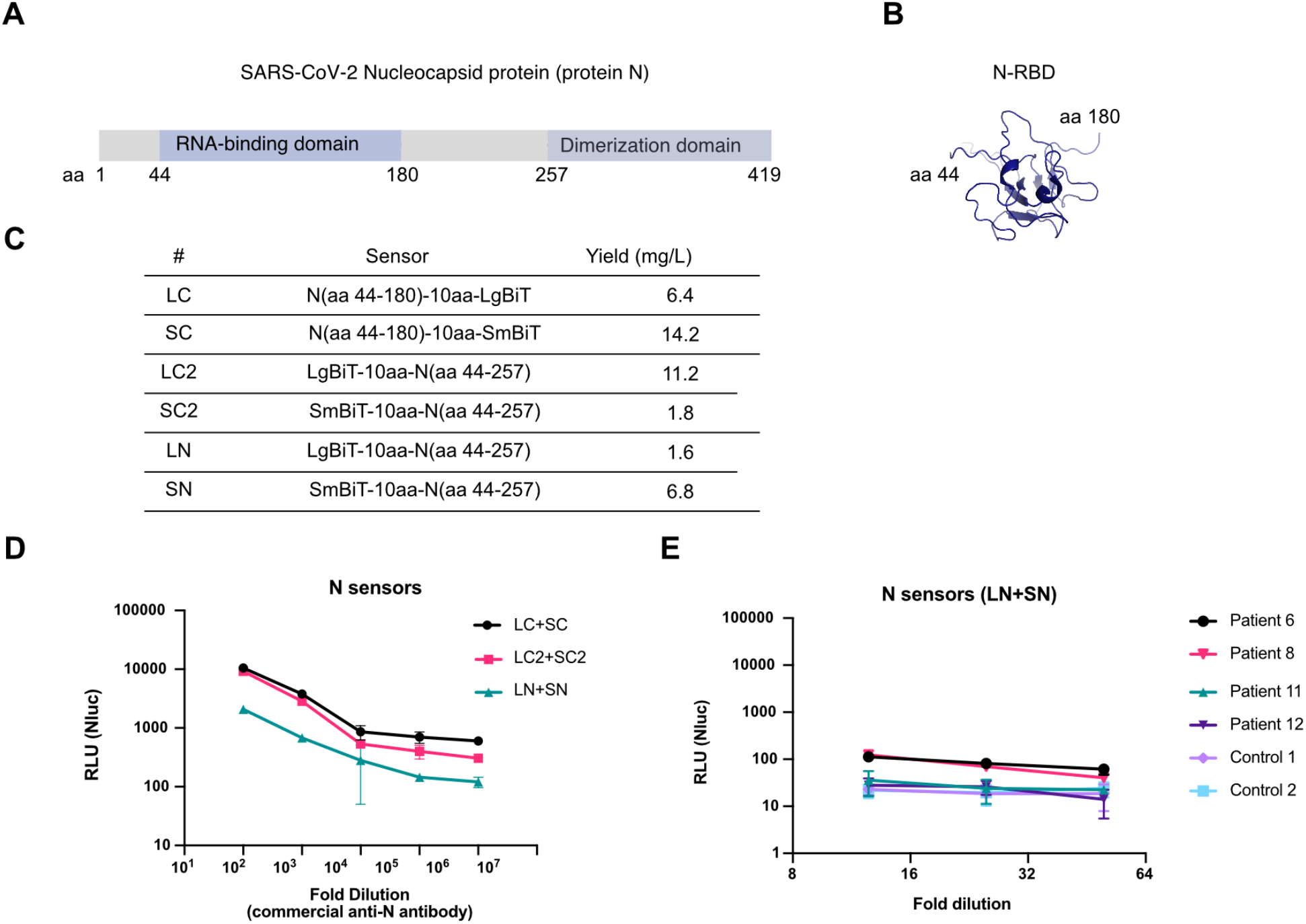
Design and characterization of N sensors. **(A)** Annotated depiction of the SARS-CoV-2 Nucleocapsid protein (protein N). All N protein fusions designed included the RNA binding domain (aa 44–180, N-RBD) and excluded the dimerization domain (aa 257–419). **(B)** Structure of the N-RBD domain shows the N and C termini locate far from each other and fusion of the split enzyme fragments to N or C termini may result in different detection sensitivity (PDB: 6YI3). **(C)** Yield of the six N protein-NanoBiT sensor fusions. **(D)** The N-terminal N sensor pair (LN + SN, 44–257) was less sensitive than the LC + SC (44–180) and LC2 + SC2 (44–257) C terminal N sensor pairs when the assay was performed on a rabbit polyclonal anti-N protein antibody (Sino Biological, Cat#: 40588-T62–50). **(E)** Additionally, only patient 6 and 8 showed signals above controls in the serological assay performed with LN + SN sensors, while all four patients showed signals with the LC + SC sensors.

**Figure S6.**
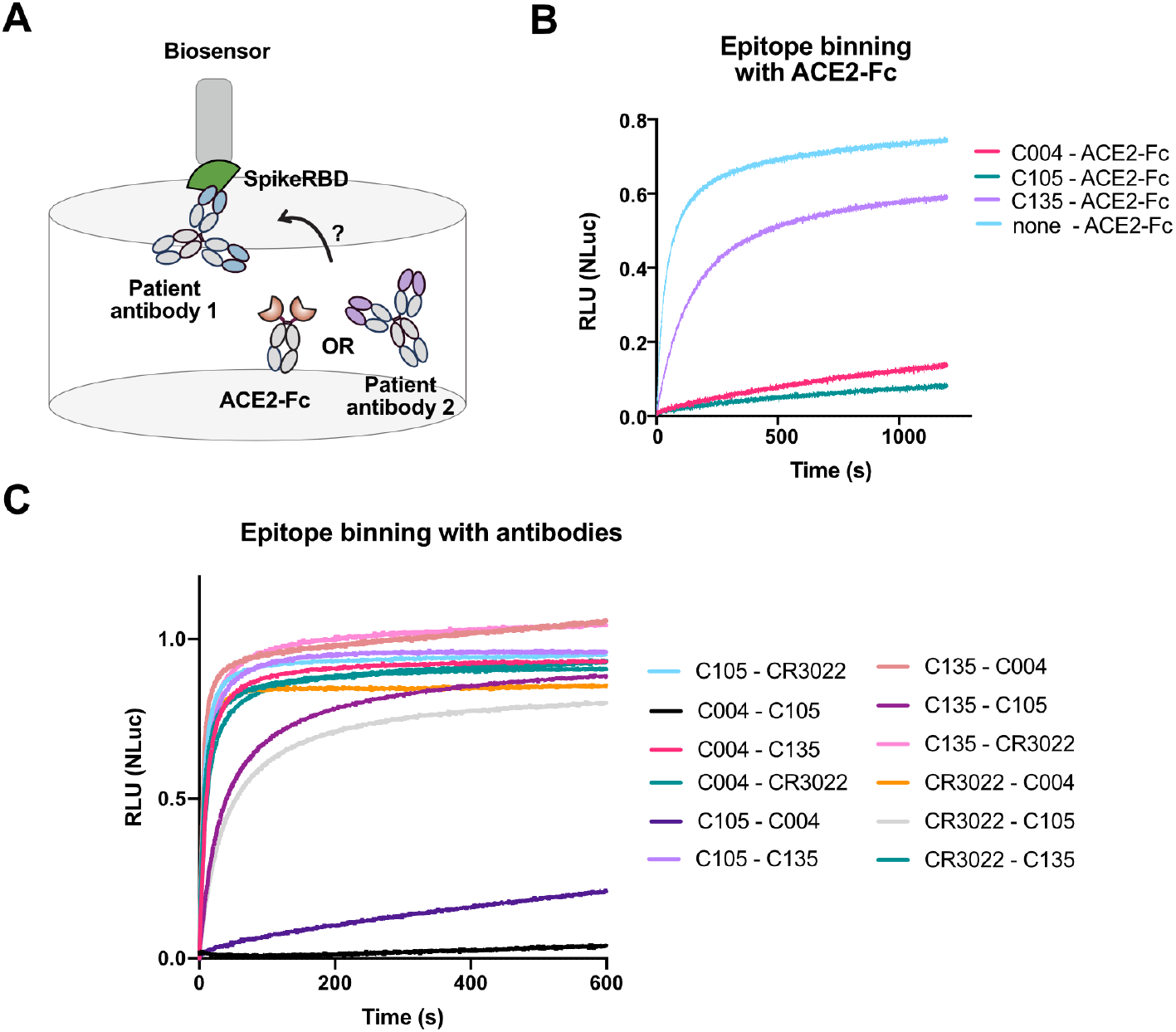
Epitope characterizations of CR3022, C004, C105 and C135. **(A)** Design of a Biolayer interferometry (BLI) experiment to characterize competitive binding of the antibodies with ACE2-Fc and other antibodies. **(B)** BLI experiments showed C004 and C105 both competed with ACE2-Fc for binding while C135 does not. **(C)** BLI experiments showed C004 competed with C105 for binding while the other antibodies do not compete.

**Figure S7.**
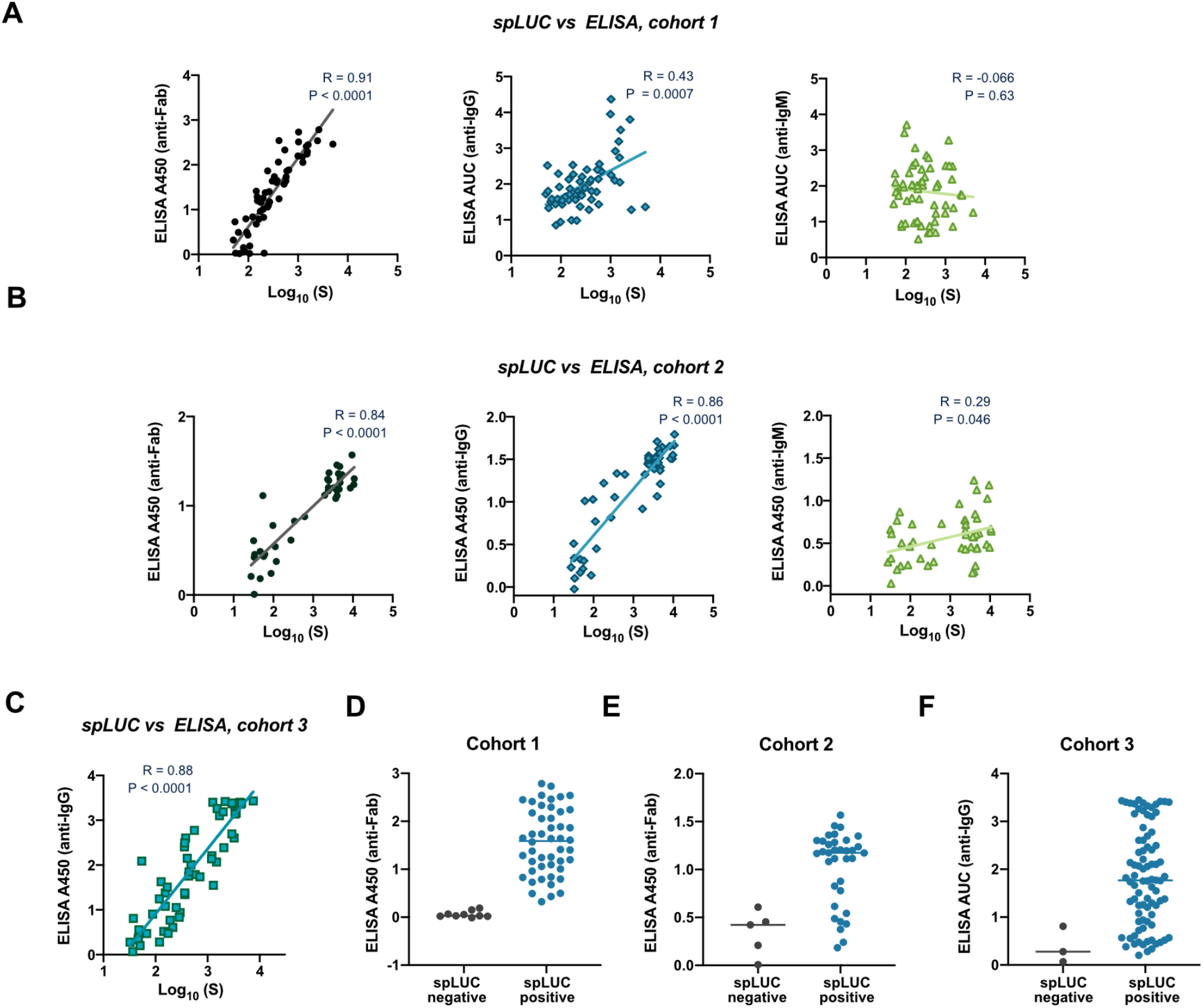
Comparison of the ELISA and the spLUC results. **(A)** Signals from the S sensor spLUC assay (cohort 1) correlate very well with S-RBD ELISA anti-Fab signals (R = 0.91), moderately well with anti-IgG signals (R = 0.43), and poorly with anti-IgM signals for cohort 1 (R = –0.066). **(B)** Signals from the S sensor spLUC assay (cohort 2) correlate very well with S-RBD ELISA anti-Fab signals (R = 0.84) and with anti-IgG signals (R = 0.86), but poorly with anti-IgM signals for cohort 1 (R = 0.29). **(C)** Signals from the S sensor (cohort 3) correlate well with S-RBD ELISA anti-IgG signals (R = 0.88). For A-C, the Spearman R values and P values are labeled in each graph. **(D-F)** The seronegative samples in the anti-S spLUC assay also showed low anti-Fab or anti-IgG signals in ELISA serology tests for cohort 1 **(D)**, cohort 2 **(E)**, and cohort 3 **(F)**.

**Figure S8.**
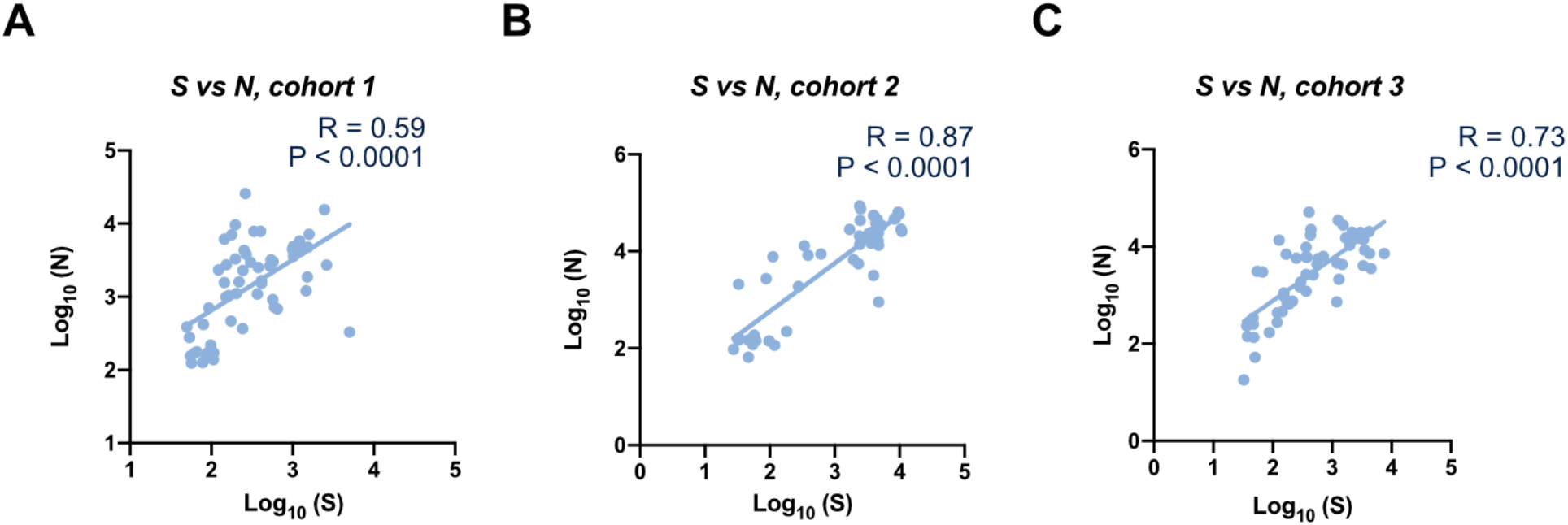
Individual cohorts show good correlation between S and N sensors. Each cohort shows robust correlation with R = 0.59, 0.87, and 0.73 for **(A)**, cohort 1, **(B)**, cohort 2, and **(C)**, cohort 3, respectively.

**Figure S9.**
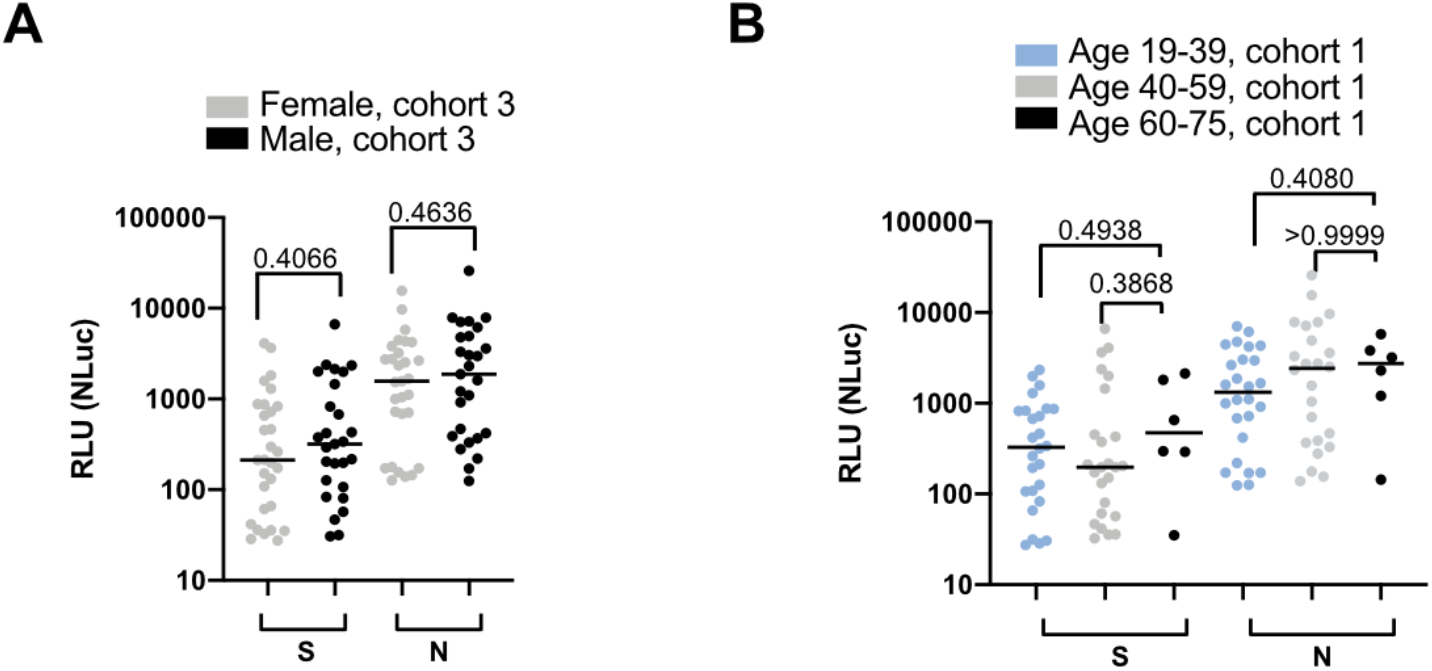
Further correlation of spLUC signal and gender/age. **(A)** For cohort 3, males show a slightly higher spLUC assay signal compared to females, although this difference is not statistically significant. **(B)** Cohort 1 spLUC signal shows no significant difference in signal among age groups.

**Figure S10.**
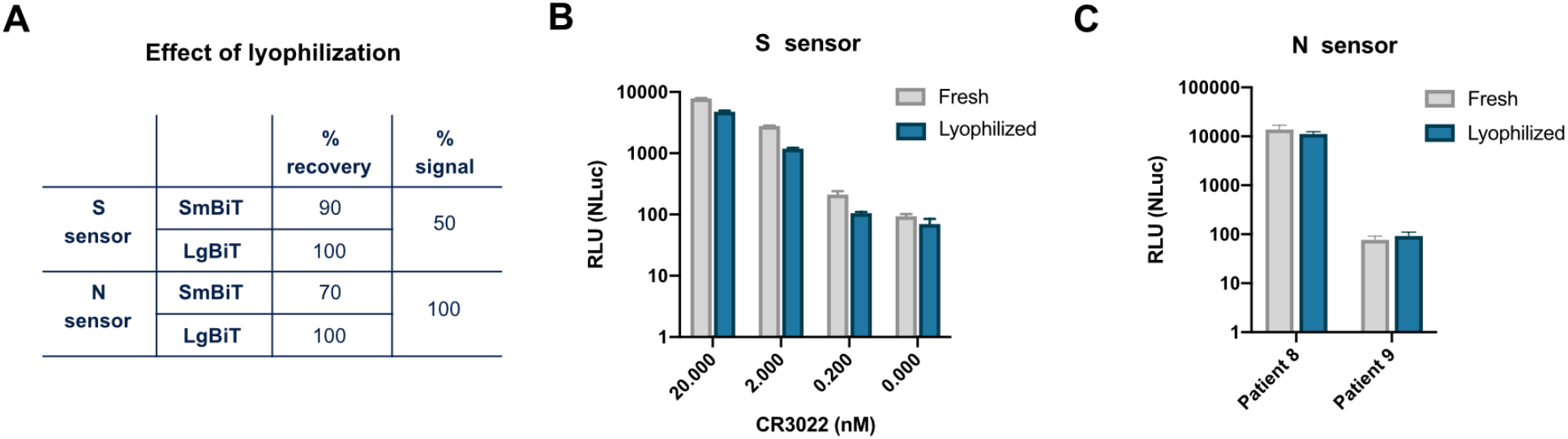
The S and N sensors were functional after lyophilization. **(A)** Both the S and the N sensors can survive lyophilization. The majority of proteins (70–100%) can be reconstituted after lyophilization. The lyophilized S sensors lost 50% of signal. The lyophilized N sensors remain 100% active. **(B)** The lyophilized S sensors detected CR3022 at ∼50% signal strength compared to fresh sensors. **(C)** The lyophilized N sensors detected antibodies from patient sera at similar signal strength compared to fresh sensors.

**Figure S11.**
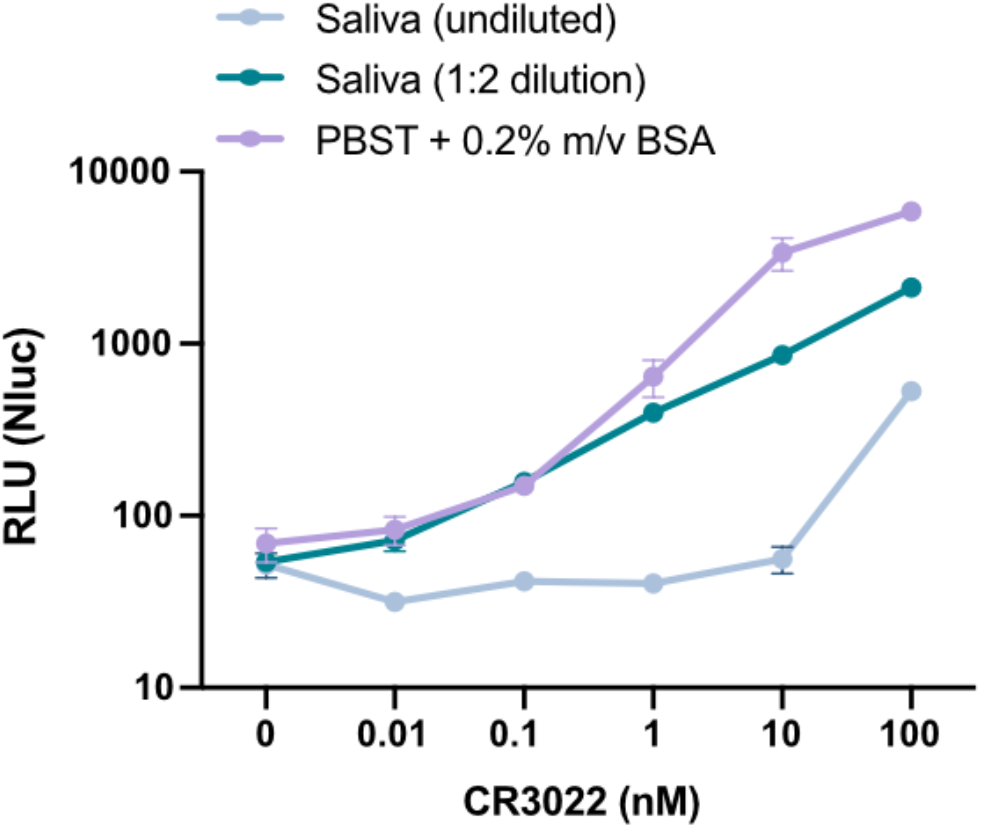
Saliva condition optimization. spLUC reactions are compatible with saliva samples. The CR3022 antibody was spiked into healthy individual saliva at 10-fold dilutions from 100 nM to 0.01 nM. While undiluted saliva reduced signal 10-fold and reduced sensitivity, 1:2 dilution of saliva only reduced signal by 3-fold and did not decrease the sensitivity. Each dot represents the average of two technical replicates and error bars represent standard deviation.

**Table S1.** Determination of assay cutoff values

|  | S | N |
| --- | --- | --- |
| SERUM DILUTIONS | 1:12.5 | 1:12.5 |
| # SAMPLES | 56 | 120 |
| MIN | 12 | 2.5 |
| MAX | 44.5 | 84 |
| MEDIAN | 23.2 | 25 |
| MEAN | 24.5 | 29.5 |
| STANDARD DEVIATION (SD) | 7.1 | 17.8 |
| DERIVED CUTOFF (MEAN+3XSD) | 45.9 | 83.1 |

